# Long-term Stored Platelets Coupled to Thrombospondin-1 Detection for Rapid and Accurate Identification of Pathogenic HIT Antibodies

**DOI:** 10.1101/2021.09.01.21262974

**Authors:** A Kanack, C Jones, B Singh, R Leger, NM Heikal, D Chen, RK Pruthi, A Padmanabhan

**Affiliations:** Department of Laboratory Medicine and Pathology, Mayo Clinic, Rochester, MN, USA; Retham Technologies, Wauwatosa, WI, USA; Department of Medicine, Mayo Clinic, Rochester, MN, USA

## Abstract

Heparin-induced thrombocytopenia (HIT) is a potentially life-threatening disorder characterized by antibodies to Platelet Factor 4 (PF4)-polyanion complexes which cause thrombocytopenia and thrombosis. Currently used technically-simple frontline assays such as the PF4-polyanion enzyme-linked immunosorbent assays (ELISAs) lack specificity, and more accurate functional assays such as the serotonin release assay (SRA) and PF4-dependent P-selectin expression assay (PEA) have long turnaround times due to technical complexity and availability only in the reference laboratory setting. There is a critical need for accurate near-patient functional testing to guide patient management, but a key barrier to attaining this goal is the short-term viability of platelets. Here, we detail a process of platelet cryopreservation that renders them viable for at least one year and show that PF4-treated cryopreserved platelets, when coupled with ELISA-based measurement of thrombospondin-1 release (a platelet **α**-granule protein), detects pathogenic HIT antibodies with high accuracy. Furthermore, testing of a cohort of non-pathogenic HIT antibodies that were strongly reactive in PF4/polyanion ELISA but negative in functional assays demonstrated negative results in the thrombospondin-1 release assay, confirming high specificity of this technique. These findings have the potential to transform the diagnostic testing paradigm in HIT by making in-hospital functional testing available for rapid and accurate diagnosis.

## Introduction

HIT is a disorder characterized by antibodies to PF4/polyanion complexes that cause thrombocytopenia and thrombosis^1- 4^. A recent large epidemiological study suggests high morbidity and a mortality rate of ∼10% in HIT patients^5^. Currently, two classes of assays are used for the HIT diagnosis. PF4/polyanion enzyme-linked immunosorbent assay (ELISA)-based tests are simple from a technical standpoint but are highly non-specific^6^. For example, in a recent 400+ patient prospective HIT diagnostic study, only ∼50% of ELISA-positive samples harbored platelet-activating antibodies^7^. Thus, reliance on these assays results in an “epidemic of overdiagnosis”^8^ and excessive use of alternative anticoagulants that have a higher risk of bleeding^6,9^. In addition to the medical risks associated with excessive use of non-heparin anticoagulants, they are expensive^10^.

On the other hand, more accurate functional (platelet-activation based) assays such as the serotonin-release assay (SRA)^11^ and heparin-induced platelet activation assay (HIPA)^12^ require complex techniques and handling of radioactive reagents or aggregometry equipment. Thus, functional assays are available only in the context of reference laboratories. Recent studies suggest that the use of PF4-treated platelets in functional HIT testing is equivalent or superior to the use of heparin-treated platelets for HIT diagnosis.^7,13,14^ A new assay developed based on these findings, the PF4-dependent P-selectin expression assay (PEA), although simpler, still requires flow cytometry^7^ and is currently offered by only one laboratory in the United States. Thus, functional testing is associated with long turnaround times, making their use impractical for immediate patient management.

Therefore, there is a critical need to develop rapid and accurate (functional) diagnostic assays for HIT that can be implemented in the near-patient, in-hospital environment. A key challenge that has prevented the development of self-contained *functional* HIT *in vitro* diagnostic assays (IVDs) is the extremely short viability of platelets. Here, we detail the development of a process of cryopreservation that extends platelet viability for at least one year. We also demonstrate that PF4-treated cryopreserved platelets can support the detection of pathogenic, platelet-activating HIT antibodies by measuring the release of an abundant platelet α-granule protein, thrombospondin-1 (TSP1) in a simple ELISA-based format.

## Methods

### Platelet Cryopreservation

Platelet-rich plasma (PRP) units sourced from whole blood donations anticoagulated with citrate phosphate dextrose from healthy blood donors were obtained two days after draw, upon completion of infectious disease testing. Anticoagulant citrate dextrose-A (1.2mM sodium citrate; 1.4mM dextrose, pH 5.9) and prostaglandin E-1 (50µg/mL stock, Millipore Sigma, St. Louis, MO) and was added 1:10 (v/v) and 1:1000 (v/v) respectively to the two day old PRP units. The PRP was centrifuged at 100g, 15 minutes. Supernatant was collected and centrifuged at 1000g 15 minutes to pellet platelets. Supernatant was discarded, and platelets were resuspended at a concentration of 1×10^6^/µL and incubated at 37°C for 2 hours in a buffer containing the cryoprotectant, trehalose (9.5 mM HEPES; 100 mM NaCl; 4.8 mM KCl; 5.0 mM glucose; 12 mM NaHCO_3_; 50 mM trehalose, pH 6.8). 30% (w/v) bovine serum albumin (BSA, Millipore Sigma, St. Louis, MO) was added to a final 4% (w/v) concentration. This platelet solution was then aliquoted in 500µL volumes per tube. Platelets were cryopreserved in a Thermo Scientific CryoMed 7450 controlled rate freezer at a range of freezing rates. Platelets cryopreserved at an optimal rate of 4°C/min to the target of -80°C are presented in this study. Platelets were stored at -80°C for various periods before testing, as noted in the *Results and Discussion* section.

### Thrombospondin Release Assay (TRA)

Thrombospondin-1 (TSP1), a highly expressed protein in platelet α-granules^15^ released upon HIT-antibody mediated platelet activation, was measured by ELISA. Briefly, after method optimization (see *Supplementary Appendix* for initial conditions), cryopreserved platelet tubes were thawed at 37°C and centrifuged at 1000xg for 5 minutes. Supernatants were discarded, and platelets were resuspended in a modified Ringer’s Citrate Dextrose (RCD) buffer containing trehalose (108mM NaCl; 3.8mM KCl; 1.7mM NaHCO_3_; 22.9M sodium citrate; 27.8mM Glucose; 50mM trehalose, pH 6.5) and centrifuged at 1000xg for 5 minutes. Supernatants were discarded. Platelets were resuspended in 80µL of a phosphate-buffered saline-based (PBS) activation buffer (137mM NaCl; 2.7mM KCl; 10mM Na_2_HPO4; 1.8mM KCl; 1% [w/v] Bovine Serum Albumin[BSA]; 50mM trehalose; 150µg/mL PF4 [Protein Foundry, Wauwatosa, WI], pH 7.4) and incubated at room temperature for 20 minutes. 20µL of patient serum was added to the PF4-treated platelets, incubated for 30 minutes at room temperature, and supernatants were collected after centrifugation at 1000xg for 5 minutes. In experiments using thrombin receptor activating peptide (TRAP), platelets were prepared similarly but were not treated with PF4. TRAP (125µg/mL stock, AnaSpec, Fremont, CA) or buffer control was added in lieu of serum to a final concentration of 25µg/mL. TSP1 was quantified from platelet supernatants using a Thrombospondin-1 DuoSet ELISA kit (R&D Systems, Minneapolis, MN) using manufacturer instructions.

### Patient Samples

Preliminary testing was performed with apheresates from therapeutic plasma exchange procedures from three patients with confirmed HIT. Thirty-four samples, 16 PEA-positive and 18 PEA-negative samples were identified among remnant sera available in adequate volumes after HIT ELISA diagnostic testing at Mayo Clinic (Rochester, MN). Two additional PEA-positive samples used were apheresates from therapeutic plasma exchange procedures on patients with confirmed HIT. All samples had IgG-specific ELISA (Immucor, Norcross, GA) and SRA results available. Studies were approved by the Institutional Review Board of Mayo Clinic.

## Results and Discussion

In initial studies, we evaluated the ability of one-week-old refrigerated (4°C) platelets to support detection of HIT antibodies in functional testing using PF4-treated platelets, compared to 2-day old platelets obtained from room temperature stored PRP bags (**Supplementary Fig S1**). Results demonstrated strong baseline platelet activation due to refrigeration, as evidenced by increased P-selectin (CD62p) expression even upon incubation with healthy donor sera (**Supplementary Fig S1**), suggesting that long-term refrigeration of platelets is not a viable option for platelet storage. Our observation of increased activation with cold-stored platelets agrees with previously published observations that refrigerated platelets undergo structural, metabolic, and functional changes that impact their activation state^16,17^. As an alternative to refrigeration, we explored the possibility of using cryopreserved platelets in functional testing. Cryopreserved platelets, developed as described in *Methods*, were initially treated with a strong platelet agonist, TRAP, which resulted in robust activation as evidenced by high TSP-1 release (190 µg/mL vs. 26 µg/mL with buffer control; **Supplementary Fig S2)**.

Next, we assessed the feasibility of using PF4-treated cryopreserved platelets to detect platelet-activating HIT antibodies. Initially, 20 platelet lots, each obtained from a different healthy donor were used in the study (**Figure 1A**). Platelets were cryopreserved as described, thawed, treated with PF4, and incubated with three platelet-activating HIT or normal control samples to assess their ability to elicit platelet activation in the thrombospondin-1 release assay (TRA). Despite some variability, HIT samples stimulated higher TSP1 release relative to controls for each of the 20 platelet lots tested (**Figure 1A**). To assess the longer-term viability of cryopreserved platelets for HIT antibody detection, the TRA was repeated using seven platelet lots that had been frozen for more than four weeks (**Figure 1B**), and demonstrated that platelets remained viable and capable of degranulation upon activation even after storage. Of note, the reactivity of platelets at these two time points was highly reproducible within each platelet lot. Together, this data suggests that while all platelets were “reactive” in the TRA, some may be more reactive than others, a phenomenon that has been documented with other HIT functional assays such as the SRA^18^. Critically, longer-term storage of platelets for 12 months supported HIT antibody-mediated, PF4-dependent platelet activation resulting in the release of significant amounts of TSP1 (**Figure 1C**).

**Figure 1.**
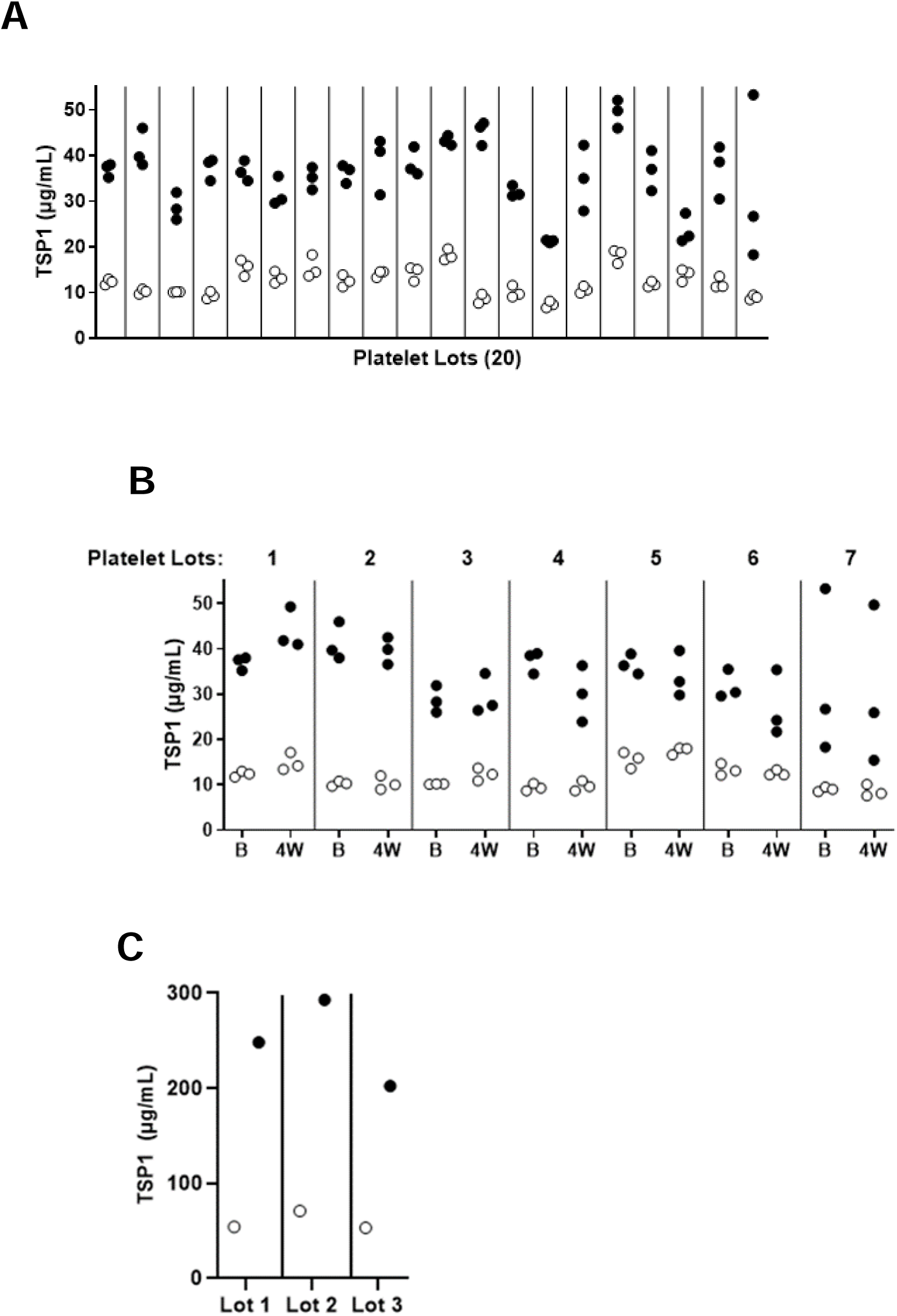
Cryopreserved frozen platelets are activated by HIT antibodies. Thrombospondin-1 release assays (TRA) was performed with three platelet-activating HIT (closed circles) and control (open circles) samples using (**A**) Twenty lots of cryopreserved platelets and (**B**) Seven cryopreserved platelet lots stored for >4 weeks at -80°C (“4W”) and compared to activation of platelets stored for ≤1 week at -80°C (“B”, baseline). (**C**) shows PF4-dependent activation induced by one HIT (closed circle) and one normal control sample (open circle) with three cryopreserved platelet lots stored at -80°C for 12-months. “4W” platelets ranged in storage age from 29 to 36 days, while “B” platelets ranged in storage age from 1 to 7 days. Date shown in (**A**) and (**B**) wee obtained using preliminary methods, as described in the *supplementary appendix*. Y-axis depicts TSP1 concentration (in µg/mL) measured in platelet supernatant and X-axis depicts platelet lot tested.

To assess the diagnostic accuracy of the cryopreserved platelet-coupled TRA, a cohort of 18 PEA-positive and 18 PEA-negative HIT-suspected patient samples were selected for the study (**Figure 2A**). The cohort was specifically selected to include several cases with “false-positive” IgG-specific ELISA results, i.e., ELISA-positive but negative in functional testing. In this cohort, PEA-SRA concordance was demonstrated in 34 of 36 samples (**Figure 2A**). Two PEA-positive samples were SRA-negative. One of the SRA-negative samples was an apheresate from a confirmed HIT-positive patient (ELISA OD 2.5, and SRA positive at 62%). The likely reason for negative SRA results with apheresate was sample dilution secondary to apheresis and is in line with published observations of lower analytical sensitivity of the SRA relative to the PEA^14^. The second PEA-positive/SRA-negative sample was from an ELISA-positive patient with thrombosis and thrombocytopenia following heparin exposure who demonstrated recovery of platelet counts upon heparin cessation, suggesting the high likelihood of SRA-negative HIT, an entity that multiple groups have recently described^2,13,19^.

**Figure 2.**
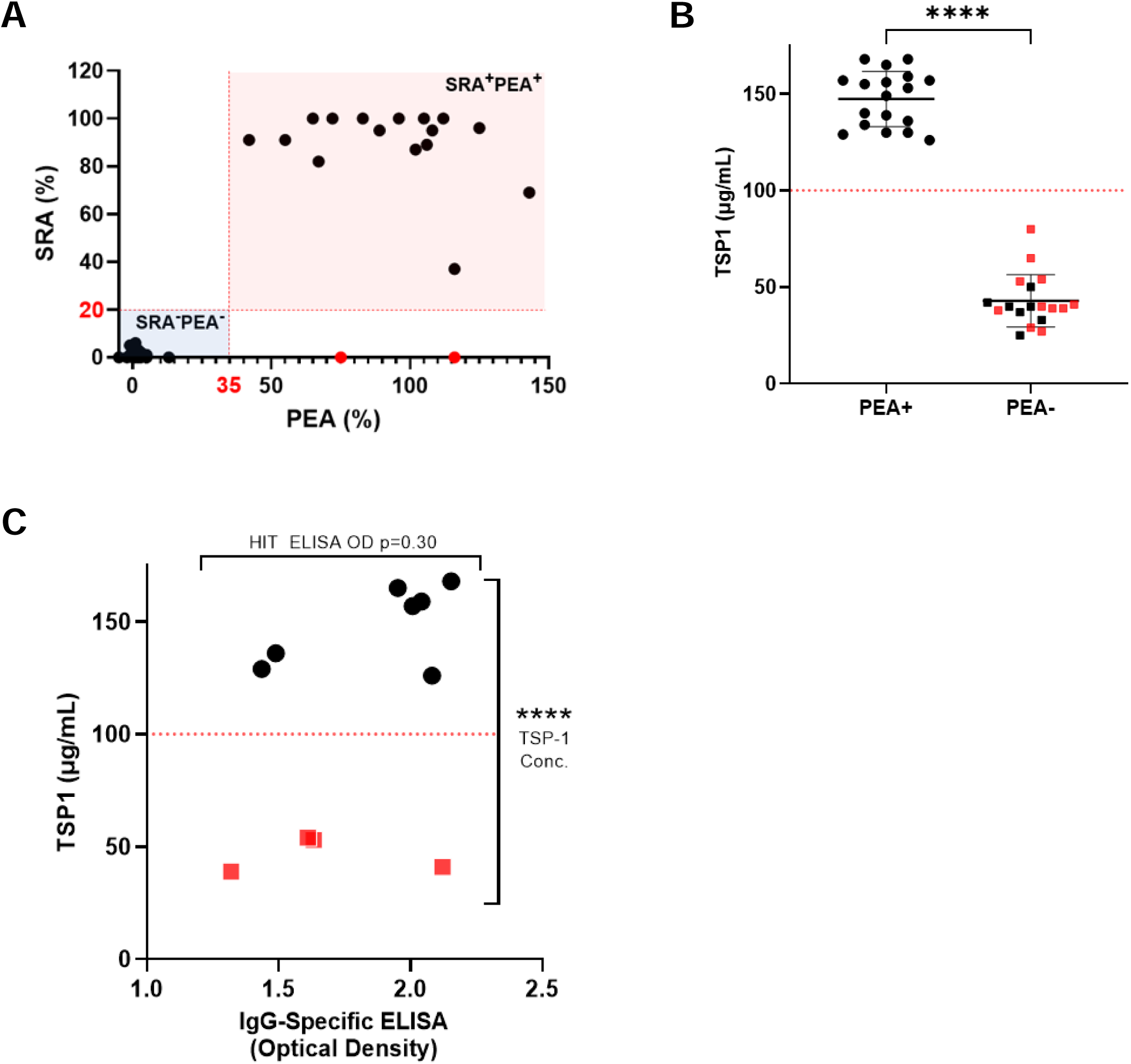
The TRA had high sensitivity and specificity for detection of pathogenic platelet-activating HIT antibodies. (A) PEA and SRA results of the patient cohort used in the study are shown on the X-axis and Y-axis, respectively. Red dotted lines represent positive cut-off values (35% p-selectin expression and 20% serotonin release). The two patient samples highlighted in red tested positive in the PEA, but negative in the SRA, and had case histories consistent with HIT. (B) TRA results are displayed for the patient cohort after qualitatively grouping based on PEA results as positive (circles), or negative (squares). Patients that tested negative in the PEA but had positive IgG-specific HIT ELISA results are displayed as red squares (11 samples). Each data point represents the mean of technical triplicates. TSP-1 concentrations (µg/mL) are depicted on the Y-axis with the mean +/-1SD for each group displayed. (C) TRA results of ELISA OD-matched PEA+/SRA+ vs PEA-/SRA-patients are presented. Seven patients had platelet-activating antibodies (closed circles) while four were did not (red squares). Y-axis depicts TSP1 concentration (in µg/mL) measured in platelet supernatant and X-axis depicts the ELISA optical density. Each data point represents the mean of technical triplicates. A two-tailed, unpaired Student’s t test was used for comparison. ****p < 0.0001.

All 18 PEA-positive samples strongly activated cryopreserved platelets, with a mean TSP1 concentration of 147 µg/mL in platelet supernatant. In comparison, the 18 non-activating PEA-negative samples induced a mean TSP1 release of 43 µg/mL and showed clear separation from the PEA-positive group (**Figure 2B**). Of note, 11 of the 18 patients in the PEA-negative cohort were ELISA-positive (**Figure 2B**). To stringently assess the specificity of the TRA, PEA-positive and PEA-negative groups matched for strong ELISA positivity (1.0-2.2 OD) from within the 36-sample cohort were compared in a subset analysis (**Figure 2C**). PEA-positive (n=7) and PEA-negative (n=4) groups had a mean ELISA O.D. of 1.88 and 1.67, respectively (p=0.30). TSP1 released by activating versus non-activating samples were significantly different with no overlap between the two groups, as shown in **Figure 2C**, demonstrating the high specificity of the cryopreserved platelet-coupled TRA. Of note, studies presented in **Figures 2B-C** were conducted using cryopreserved platelets that were shipped on dry ice for ∼30 hours, demonstrating the viability of platelets with cold chain-compatible transport. Proof-of-concept studies presented in **Supplemental Figure S3** also demonstrate that heparin-treated cryopreserved platelets, like PF4-treated platelets, can be used to detect platelet-activating HIT antibodies.

In summary, using a heterogeneous group of patient samples, including those that were strongly false-positive in the ELISA, cryopreserved platelets coupled to a TSP-1 ELISA endpoint demonstrated 100% sensitivity and specificity for the detection of platelet-activating HIT antibodies. While platelet activatability after long-term storage (12-months) has been demonstrated, studies to evaluate platelet stability at 18 and 24-months are ongoing. The ability to use 2-day old platelets from PRP units, which are available in abundance from the blood banking industry, should facilitate large-scale production of platelets for *in vitro* diagnostic kit use. Pooling of platelets, a strategy that we have used successfully for a recently deployed functional HIT diagnostic assay, the PEA^7^, will be employed in this setting to minimize lot variation in platelet activatability. These findings have the potential to transform the diagnostic testing paradigm in HIT by making in-hospital functional testing available for rapid and accurate diagnosis by the deployment of a self-contained HIT IVD assay that will not require the user to obtain donor platelets. Emerging data suggest that anti-PF4 antibodies in Vaccine-induced immune thrombocytopenia and thrombosis (VITT) seen after some COVID-19 vaccines are consistently detected using PF4-treated platelets as opposed to heparin-treated platelets^20-22^. Thus, findings reported here may be relevant for the detection of VITT antibodies, and studies are planned.

## Data Availability

Data presented in the manuscript with be shared with legitimate parties, upon reasonable request.

## Acknowledgments

We would like to thank Stephanie Hafner, BS from Mayo Clinic’s Research Innovation Office for exceptional research coordination support. This work was supported, in part, by National Institutes of Health grant HL147734 (C.J), HL133479 (A.P) and through a grant from the Therapeutic Accelerator Program of the Drug Discovery Center at Medical College of Wisconsin and Wisconsin Economic Development Corporation’s (WEDC): Targeted Industry projects.

## Authorship

AK, CJ, BS provided critical input on laboratory and clinical elements of the study. AK and CJ performed the Thrombospondin-1 release assay and flow cytometry studies. RL, NMH, DC, and RKP provided helpful advice on clinical and laboratory aspects of the manuscript. AP conceived the experimental plan and directed the laboratory studies. AK, CJ, and AP wrote the first draft and all authors provided input and approved the final version.

