## Supplementary Appendix for "Long-term Stored Platelets Coupled to Thrombospondin-1 Detection for Rapid and Accurate Identification of Pathogenic HIT Antibodies"

Mayo Clinic

Rochester, MN 55906

**Supplementary Figures**

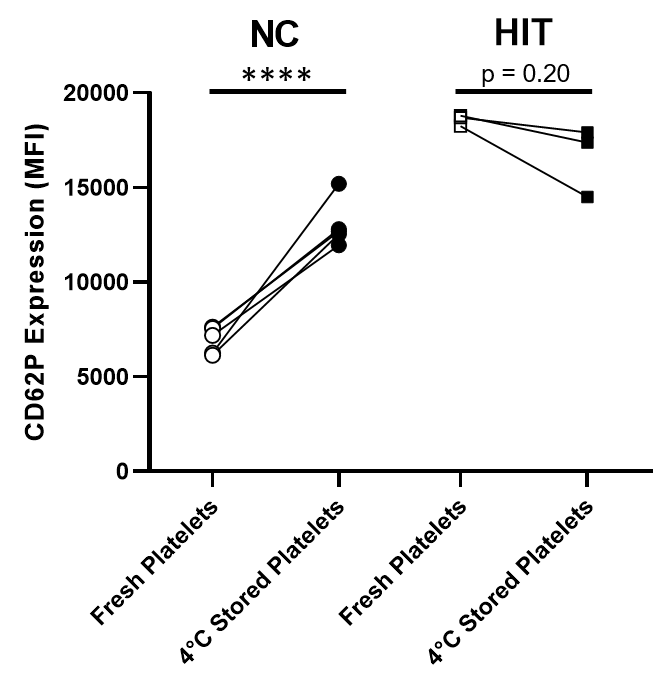

**Supplemental Figure 1. Refrigeration of platelets increases baseline platelet activation.** P-selectin expression levels were quantified by flow cytometry after incubation of five normal control (NC; circles) or three platelet-activating HIT sera (HIT; squares) with PF4-treated platelets as previously described^1^. The lines connect results from “fresh” platelets (2-day old, room temperature-stored; open symbols) with those obtained using 1-week old refrigerated platelets (closed symbols). For refrigerated platelet studies, 2-day old platelets were stored at 4°C for an additional five days and tested in the assay when platelets were seven days old. The Y-axis depicts p-selectin (CD62P) median fluorescence intensity (MFI), and the X-axis shows platelet storage conditions. Each data point represents the mean value of technical duplicates, and groups are compared using two-tailed, unpaired Student’s t test (NC) or two-tailed, unpaired t test with Welch’s correction (HIT). ****P < 0.0001.

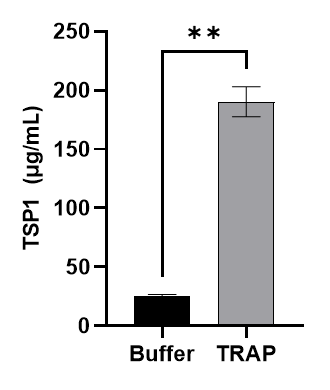

**Supplemental Figure 2. Thrombin receptor activating peptide (TRAP) activates cryopreserved platelets.** Cryopreserved platelets were thawed, washed, and incubated with TRAP (25 µg/mL, final concentration) for 30 minutes, and TSP-1 concentrations were quantified in the platelet supernatant using a TSP-1 ELISA. Y-axis depicts TSP1 concentration (µg/mL). Data are from technical triplicates with means ± 1 SD are displayed. Results are compared using two-tailed, unpaired t test with Welch’s correction. **P < 0.01.

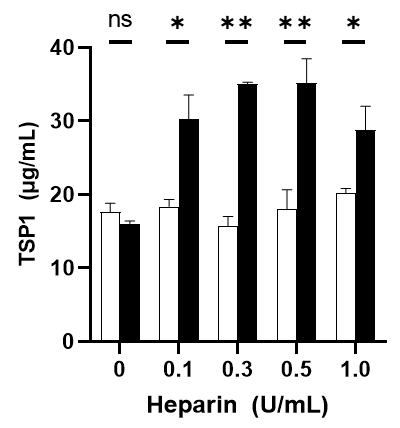

**Supplemental Figure 3. HIT antibodies activate heparin-treated cryopreserved platelets**. Cryopreserved platelets were thawed, washed, treated with unfractionated heparin followed by incubated with one normal control (open bar) or platelet-activating HIT (closed bar) sample. TSP-1 concentration in platelet supernatant was quantified. Y-axis depicts TSP-1 concentration (µg/mL), and X-axis shows the final heparin concentration in the reaction (U/mL). Data are from technical triplicates with means ± 1 SD displayed. Comparison was performed using two-tailed, unpaired t test with Welch’s correction. *P < 0.05, **P < 0.01, ns=not significant.

**Supplementary Methods**

In preliminary experiments (presented in **Figs 1A, 1B** and **Supplementary Figure S3**), platelets obtained from healthy donors, as noted above, were cryopreserved in 1000µL volumes, thawed at 37°C and then centrifuged at 1000xg for 15 minutes. Supernatants were discarded and platelets were resuspended in a modified RCD buffer (108mM NaCl; 3.8mM KCl; 1.7mM NaHCO_3_; 22.9M sodium citrate; 27.8mM Glucose, pH 6.5) and centrifuged at 1000xg for 15 minutes. Supernatants were discarded. Platelets were resuspended in 175µL of a PBS-based buffer (137mM NaCl; 2.7mM KCl; 10mM Na_2_HPO4; 1.8mM KCl; 1% [w/v] BSA, pH 7.4) and aliquots were pooled. 97.5µL of PF4 (800µg/mL stock) was then added to 422.5 µL of the platelet suspension. Alternatively, 25µL of heparin (Millipore Sigma, St. Louis, MO) at 0 (buffer control), 1, 3, 5, and 10U/mL stock concentration were added to 175µL of the platelets suspension and incubated for 20 minutes. 20µL of patient serum was then added to 80µL of PF4 or heparin treated platelets resulting in a final PF4 concentration of 120µg/mL or heparin concentrations of 0.1U/mL, 0.3U/mL, 0.5U/mL or 1.0U/mL. This mixture was incubated for 30 minutes at room temperature. Supernatants were collected after centrifugation at 1000xg for 15 minutes. Cryopreserved platelets were also lyophilized but were not activatable by a strong platelet agonist (Thrombin receptor-activating peptide, TRAP) or HIT samples (data not shown).
